# Antimicrobial resistance trends from a hospital and diagnostic facility in Lahore, Pakistan: A five-year retrospective analysis (2014-2018)

**DOI:** 10.1101/19012617

**Authors:** Carly Ching, Summiya Nizamuddin, Farah Rasheed, R.J. Seager, Felix Litvak, Faisal Sultan, Muhammad Zaman

## Abstract

**Objective:** Observing antimicrobial resistance (AMR) trends is critical to identify emerging pathogens and potential disease outbreaks. Determining these trends also allows for policy evaluations and development of interventions. We performed a retrospective analysis of microbiological testing results from a hospital and diagnostic facility in Lahore, Pakistan that represents country-wide sampling. Within this analysis, data was disaggregated by nationality, as it has been suggested that migration increases the burden of AMR. We sought to determine any trends in AMR among populations, which are often at-risk, while contributing to AMR surveillance in Pakistan, which currently does not have a national surveillance network.

**Methods:** Retrospective analysis of antimicrobial susceptibility records from 2014 to 2018 from Shaukat Khanum Memorial Cancer Hospital & Research Centre (SKMCH&RC) in Lahore, Pakistan was performed. All positive microbiological cultures from patients was analyzed to assess antibiotic resistance rates of the most common bacterial isolates and incidence of ESKAPE pathogens and emerging outbreaks among adults and children.

**Results:** For all years, data for a total of 12,702 and 78,130 bacterial and fungal isolates from children and adults, respectively, with Pakistani nationality were analyzed. For all years, data for 597 and 2470 bacterial and fungal isolates for children and adults, respectively, with Afghan nationality were analyzed. AMR rates largely did not vary between populations, but rather followed similar trends. AMR rates also largely agreed with the World Health Organization Global Antimicrobial Surveillance System results for Pakistan.

**Conclusion:** Pakistan requires increased AMR surveillance to identify emerging resistance infections and outbreaks.

## 1. Introduction

Antimicrobial resistance (AMR) is a major health concern worldwide. As of 2018, Pakistan was the 5^th^ most populous country in the world with 212.2 million people [1]. While Pakistan has a national action plan to combat AMR[2], there is no national surveillance network, and global reporting is done through the World Health Organization Global Antimicrobial Surveillance System (GLASS) [3]. However, there is a voluntary coalition from major hospitals and institutions that self-report antimicrobial resistance data, known as the Pakistan Antimicrobial Resistance Network (PARN). A recent national situational report from 2018 highlighted the burden and challenges of antimicrobial resistance in Pakistan[4]. Indeed, it has been recognized as a national concern since 2014, however without comprehensive surveillance data it is difficult to make accurate estimates of the burden of AMR throughout the country.

Adding to the complex situation in Pakistan is the influx of migrants and displaced populations. The United Nations refugee agency estimates that Pakistan hosts more than 1.4 million registered Afghan refugees [5]. It has been postulated that migrant groups add to the burden of AMR due to poor sanitation conditions and access to medicines and healthcare, however this sentiment is often anecdotal or without supporting data which can cause unnecessary stigmas. A recent comprehensive systematic review that looked at the pooled prevalence of AMR among migrant groups in Europe found that AMR was higher in refugees and asylum seekers than other migrant groups, but did not find evidence of higher rates of transmission of AMR from migrant to host populations [6].

Here, we add to AMR surveillance data in Pakistan, broken down by nationality (either Pakistani or Afghan nationals). The strength of this data set is the large sample size, much larger than GLASS reporting sample sizes for Pakistan, and the ability to analyze temporal trends. Limitations include that official migrant status of patients and patient medical history is unknown, therefore interpretation of data should be done with caution.

## 2. Methods

### 2.1 Sample collection

The retrospective descriptive study was conducted at Shaukat Khanum Memorial Cancer Hospital & Research Centre (SKMCH&RC), Lahore, Pakistan, and comprised data related to all microbiological cultures reported positive between 2014 and 2018. The hospital is a 195-bed non-profit tertiary-care specialist cancer hospital with a referral base from all over the country and adjoining regions. Additionally, the laboratory attached with the hospital runs through a network of laboratory collection centers located in 50 major cities and towns of Pakistan. All data was retrieved from the online records on the in-house information system database.

### 2.2 Antimicrobial susceptibility testing/Bacterial identification

All bacterial isolates tested during the above time period were identified by standard techniques and antimicrobial susceptibility testing was performed and interpreted according to Clinical Laboratory Standards Institute (CLSI) criteria.

### 2.3 Data Extraction

Migrants are defined as individuals born outside Pakistan; however, their patient history and migrant status is unknown. Children were defined as individuals under the age of 18. Data was extracted from de-identified patient records stored in a standardized Excel format using a purpose-built Python (version 3.7.3) script employing the Pandas package for data extraction and analysis. Script is available upon request. Data for only registered patients has a low sample size and is available upon request.

## 3. Results

### 3.1 Study Population

Among all years, data for a total of 12 702 and 78 130 bacterial and fungal isolates from children and adults, respectively, with Pakistani nationality were obtained. Among all years, data for 597 and 2470 bacterial and fungal isolates for children and adults, respectively, with Afghan nationality were obtained (Table 1).

**Table 1.** Characteristics of Study Population, 2014-2018.

| **Nationality : Pakistan**<br>Identified bacterial isolates, n |  |  |  |  |  |  |
| --- | --- | --- | --- | --- | --- | --- |
| **Year** | **Children** |  |  | **Adults** |  |  |
|  | Male | Female | Total | Male | Female | Total |
| 2014 | 1375 | 1074 | 2449 | 7511 | 6970 | 14481 |
| 2015 | 1092 | 932 | 2024 | 7000 | 6753 | 13753 |
| 2016 | 1341 | 1058 | 2399 | 7541 | 7737 | 15278 |
| 2017 | 1408 | 1383 | 2791 | 8188 | 8891 | 17079 |
| 2018 | 1594 | 1445 | 3039 | 8472 | 9067 | 17539 |
| Total | 6810 | 5892 | 12 702 | 38 712 | 39 418 | 78 130 |
| **Nationality: Afghanistan**<br>Identified bacterial isolates, n |  |  |  |  |  |  |
| **Year** | **Children** |  |  | **Adults** |  |  |
|  | Male | Female | Total | Male | Female | Total |
| 2014 | 54 | 8 | 62 | 164 | 165 | 329 |
| 2015 | 66 | 69 | 135 | 253 | 154 | 407 |
| 2016 | 73 | 54 | 127 | 332 | 244 | 576 |
| 2017 | 106 | 31 | 137 | 305 | 206 | 511 |
| 2018 | 94 | 42 | 136 | 371 | 285 | 656 |
| Total | 393 | 204 | 597 | 1425 | 1054 | 2479 |

### 3.2 Distribution of bacterial isolates

Among the top 5 most common bacteria isolated among adults for both populations were *Escherichia coli, Staphylococcus aureus, Pseudomonas aeruginosa* and *Klebsiella pneumoniae*, and this did not change between 2014 to 2018 (Table 2). In the adult Pakistani population *Enterococcus* species filled out the top 5 most common bacterial isolate for each year, while for the Afghan population it was *Staphylococcus epidermidis*. For Pakistani children, a similar distribution including *S. epidermidis* is seen. For Afghan children, *S. epidermidis, E. coli* and *S. aureus* are the most common bacteria across all years (Table 3). The full distribution of bacterial infections for each year is provided in Supplemental File 1.

**Table 2.** Top 5 Most Frequent bacteria isolated in adults, 2014-2018.

| Nationality: Pakistan |  |  | Nationality: Afghanistan |  |  |
| --- | --- | --- | --- | --- | --- |
|  | Bacteria | Adults, n (%) |  | Bacteria | Adults, n (%) |
| 2014 | *Escherichia coli* | 4475(30.9) | 2014 | *Pseudomonas aeruginosa* | 49 (14.9) |
|  | *Staphylococcus aureus* | 2261(15.6) |  | *Escherichia coli* | 49 (14.9) |
|  | *Pseudomonas aeruginosa* | 1555(10.7) |  | *Staphylococcus aureus* | 48 (14.6) |
|  | *Enterococcus species* | 892(6.2) |  | *Klebsiella pneumoniae* | 31 (9.4) |
|  | *Klebsiella pneumoniae* | 885(6.1) |  | *Staphylococcus epidermidis* | 22 (6.7) |
|  | *Other* | 4413(30.5) |  | *Other* | 130(39.5) |
| 2015 | *Escherichia coli* | 4556(33.1) | 2015 | *Escherichia coli* | 85 (20.9) |
|  | *Staphylococcus aureus* | 1907(13.9) |  | *Staphylococcus aureus* | 78 (19.2) |
|  | *Pseudomonas aeruginosa* | 1447(10.5) |  | *Pseudomonas aeruginosa* | 63 (15.5) |
|  | *Enterococcus species* | 1060(7.7) |  | *Klebsiella pneumoniae* | 31 (7.6) |
|  | *Klebsiella pneumoniae* | 974(7.1) |  | *Staphylococcus epidermidis* | 28(6.9) |
|  | *Other* | 3809(27.7) |  | *Other* | 122(30.0) |
| 2016 | *Escherichia coli* | 5075(33.2) | 2016 | *Staphylococcus aureus* | 142(24.7) |
|  | *Staphylococcus aureus* | 2078(13.6) |  | *Escherichia coli* | 100(17.4) |
|  | *Pseudomonas aeruginosa* | 1468(9.6) |  | *Pseudomonas aeruginosa* | 77(13.4) |
|  | *Klebsiella pneumoniae* | 1154(7.6) |  | *Staphylococcus epidermidis* | 51(8.9) |
|  | *Enterococcus species* | 1050(6.9) |  | *Klebsiella pneumoniae* | 43(7.5) |
|  | *Other* | 4453(29.1) |  | *Other* | 163(28.3) |
| 2017 | *Escherichia coli* | 5602(32.8) | 2017 | *Staphylococcus aureus* | 115(22.5) |
|  | *Staphylococcus aureus* | 2445(14.3) |  | *Escherichia coli* | 98(19.2) |
|  | *Pseudomonas aeruginosa* | 1594(9.3) |  | *Pseudomonas aeruginosa* | 62(12.1) |
|  | *Klebsiella pneumoniae* | 1298(7.6) |  | *Staphylococcus epidermidis* | 37(7.2) |
|  | *Enterococcus species* | 1244(7.3) |  | *Klebsiella pneumoniae* | 30(5.9) |
|  | *Other* | 4896(28.7) |  | *Other* | 169(33.1) |
| 2018 | *Escherichia coli* | 5888(33.6) | 2018 | *Staphylococcus aureus* | 170(25.9) |
|  | *Staphylococcus aureus* | 2384(13.6) |  | *Escherichia coli* | 118(18.0) |
|  | *Pseudomonas aeruginosa* | 1559(8.9) |  | *Pseudomonas aeruginosa* | 88(13.4) |
|  | *Klebsiella pneumoniae* | 1321(7.5) |  | *Staphylococcus epidermidis* | 42(6.4) |
|  | *Enterococcus species* | 1255(7.2) |  | *Klebsiella pneumoniae* | 41(6.3) |
|  | *Other* | 5132(29.3) |  | *Other* | 197(30.0) |

n=number of isolates, % refers to % of all isolates

**Table 3.**
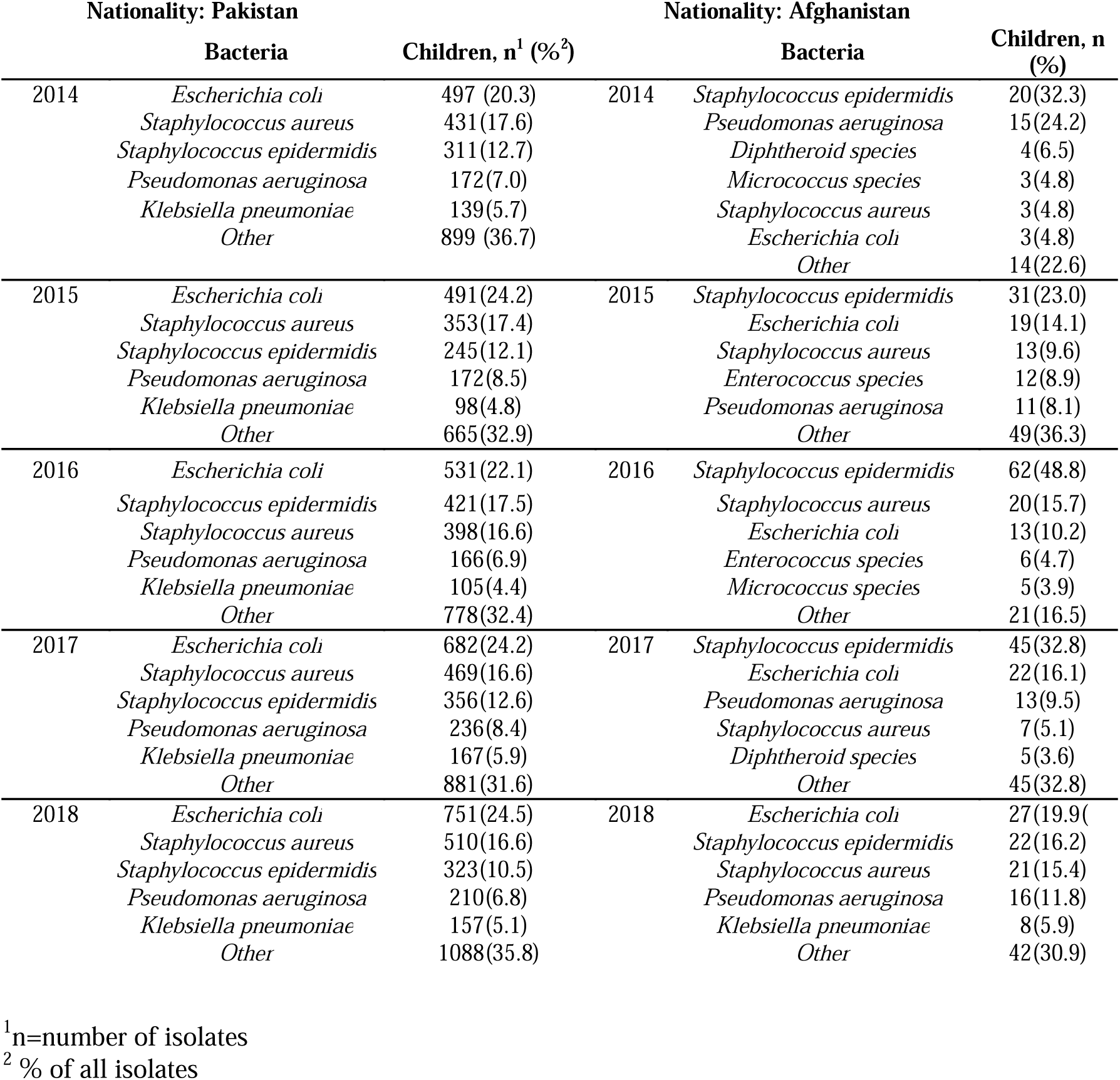
Top 5 Most Frequent bacteria isolated in children, 2014-2018.

### 3.3 Trends in incidence of ESKAPE pathogens and Salmonella typhi

The ESKAPE pathogens are the leading cause of hospital acquired infections[7], and as such we mapped the trends in incidence of these bacteria from 2014 to 2018. *S. aureus* is the most prevalent ESKAPE pathogen in adults for both populations, followed by *P. aeruginosa* and *K. pneumoniae* (Fig. 1A&B). This does not change between 2014 and 2018. Plotting this data for children from Pakistan shows a similar trend and rates to adult (Fig. 1C). Similar analysis was not performed for Afghanistan due to low numbers (Table S4). The incidence of *Enterococcus faecium, Acinetobacter baumannii* and *Enterobacter* spp. remain low.

**Figure 1.**
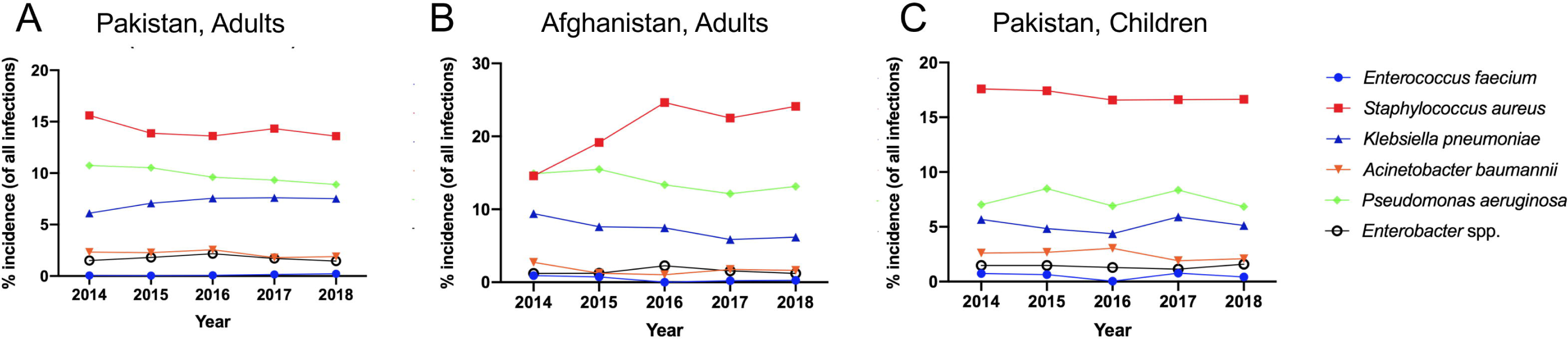
Trends in incidence of ESKAPE pathogens isolated from 2014-2018. Incidence of ESKAPE pathogens from clinical isolates from SKCM&RC from 2014-2018

While the incidence of *A. baumannii* did not increase dramatically (Fig. 1), we wanted to determine if resistance to last-line antibiotics, carbapenem and colistin, changed. Carbapenem resistant *A. baumannii* is an emerging infection in Pakistan[8]. We find that resistance to imipenem and meropenem for both adults and children from Pakistan are within the reported confidence intervals for data reported to GLASS in 2017 (53-72%)[9] and appear to be on a downward trend from 2014 to 2018 (75.4% resistant to imipenem in 2014, 66.6% in 2018). Colistin resistance in *A. baumannii* remains low ((highest percentage was 1.7% reported in 2016, Table 4). For the population from Afghanistan, there were less than ten *A. baumannii* isolates, so resistance rates were not calculated

**Table 4.**
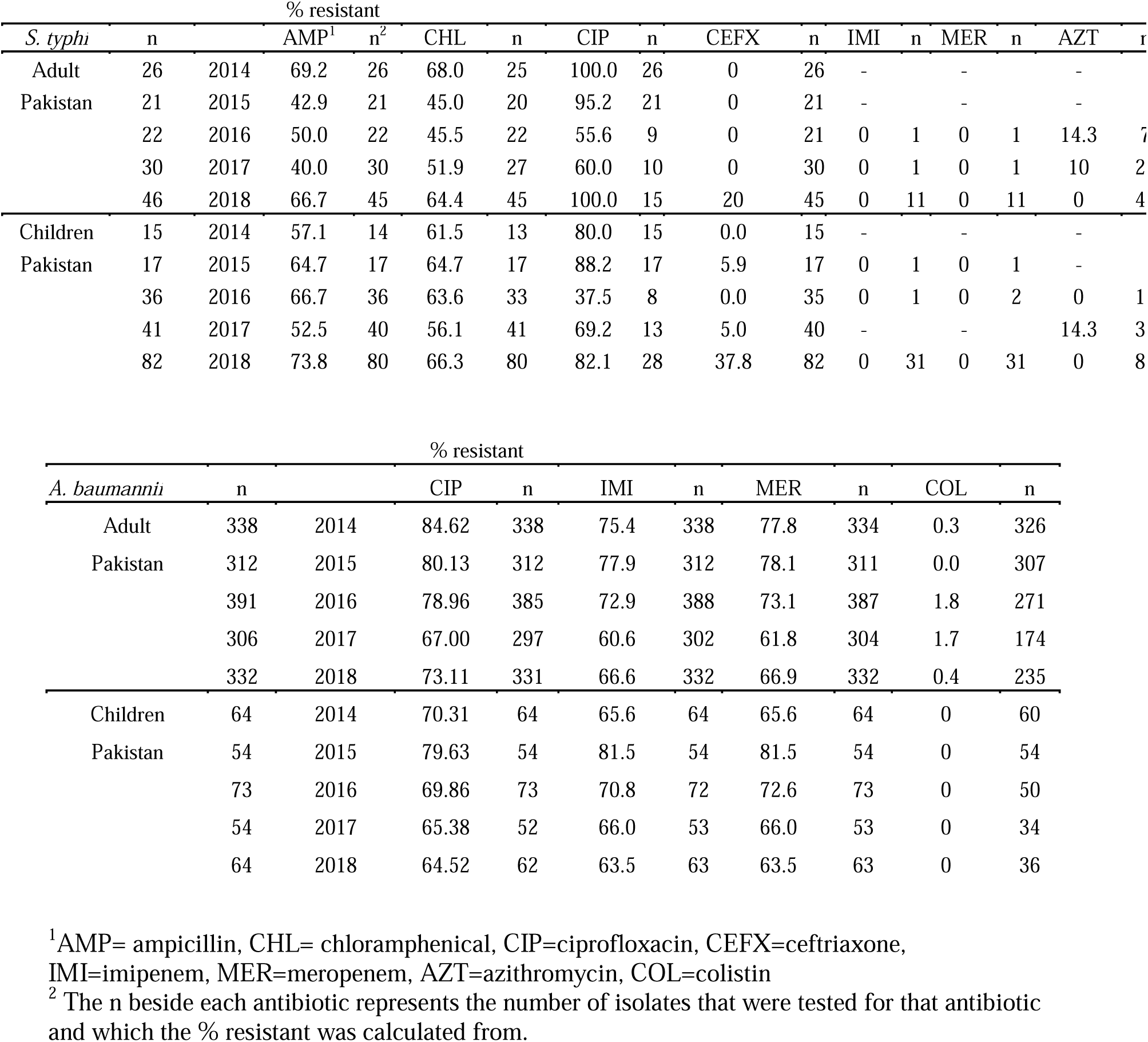
Selected Antibiotic Resistance Profile for *Salmonella typhi and Acinetobacter baumannii*, 2014-2018.

We also looked at the incidence of multidrug-resistant *Salmonella typhi* which have recently occurred in Pakistan. Since 2016, over 300 extensively drug resistant (XDR) typhoid cases emerged in Sindh, Pakistan. These bacteria were resistant to fluoroquinolones and the third-generation cephalosporin ceftriaxone but typically still susceptible to azithromycin and carbapenems [10]. Among the adult population from Pakistan, from 2014 to 2017 there were no cases of ceftriaxone resistant *S. typhi*, however in 2018, 20% of cases (n=45) were now ceftriaxone resistant (Table 4). No resistant to carbapenems and low resistance to azithromycin were observed (Table 4). In Pakistani children, low resistance to ceftriaxone was observed (0-5.9%) between 2014 to 2017. In 2018, 37.8% of *S. typhi* isolates (n=82) were resistant to ceftriaxone but none were resistant to azithromycin. In 2017 however, 14.3% (n=35) of *S. typhi* isolates from Pakistani children tested against azithromycin were resistant. There were no detected cases of *S. typhi* in Afghan adults and only 1 case in 2018 in children which was sensitive to all antibiotics tested.

### 3.4 Antibiotic resistance profiles and temporal trends

We extracted the antimicrobial susceptibility data of the most common bacterial isolates *Escherichia coli, Staphylococcus aureus, Pseudomonas aeruginosa* and *Klebsiella pneumoniae* for each year. The full table of resistance of these bacterial is provided in the supplemental data (Supplemental Tables S1-S4).

For adults, we compared trends in % resistant for fluoroquinolones and carbapenems between the two populations. We also looked at the % of oxacillin resistant *S. aureus* (MRSA) isolates between the adult populations. Since the sample size for children from Afghanistan were very low (below 10 and 20) we did not perform a comparative analysis. Looking at ciprofloxacin, % resistance is similar between the two population, with the Afghan resistant rates being lower than the population from Pakistan (Fig. 2), except for 2015 and 2016 for *E. coli* in which ciprofloxacin resistance increases before decreasing (Fig. 2A). Interestingly, for *S. aureus* we see that the upward trend follows each other (Fig. 2B). For imipenem, resistance is overall low for both populations (Fig. 3). Again for *P. aeruginosa* and *K. pneumoniae* resistance rates for the Afghan population are overall lower than that of Pakistan. In *K. pneumoniae*, resistance to carbapenems seems to be increasing for both populations (Fig. 3C). For *P. aeruginosa* for both ciprofloxacin and imipenem, the % resistant for Pakistan appears to be stable while the % resistant for the Afghan population is trending upwards but has not reached the levels of isolates of those from Pakistan (Fig. 2C & 3B). For oxacillin resistance for *S. aureus* and *S. epidermidis*, rates are very similar and strikingly follow a very similar trend (Fig. 4). It should also be noted, that the sample size between the Pakistani and Afghan population differed by over 25-fold which makes direct comparison complicated.

**Figure 2.**
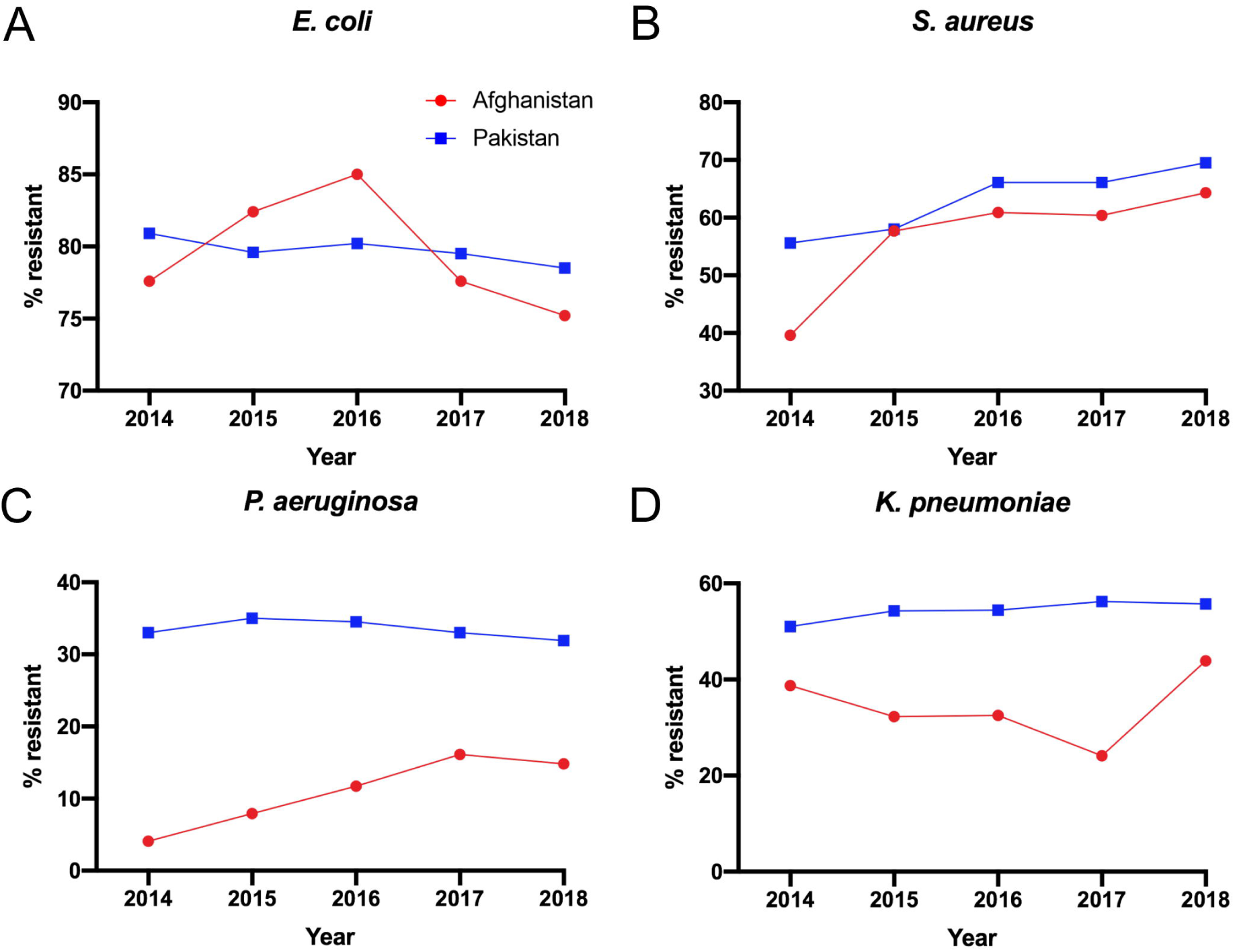
Trends in ciprofloxacin resistance in Adult patients, 2014-2018. Trends in resistance to ciprofloxacin for (A) *E. coli*, (B) *S. aureus*, (C) *P. aeruginosa* and (D) *K. pneumoniae* isolates from adult populations with nationality from Pakistan or Afghanistan taken from SKCM&RC, from 2014-2018

**Figure 3.**
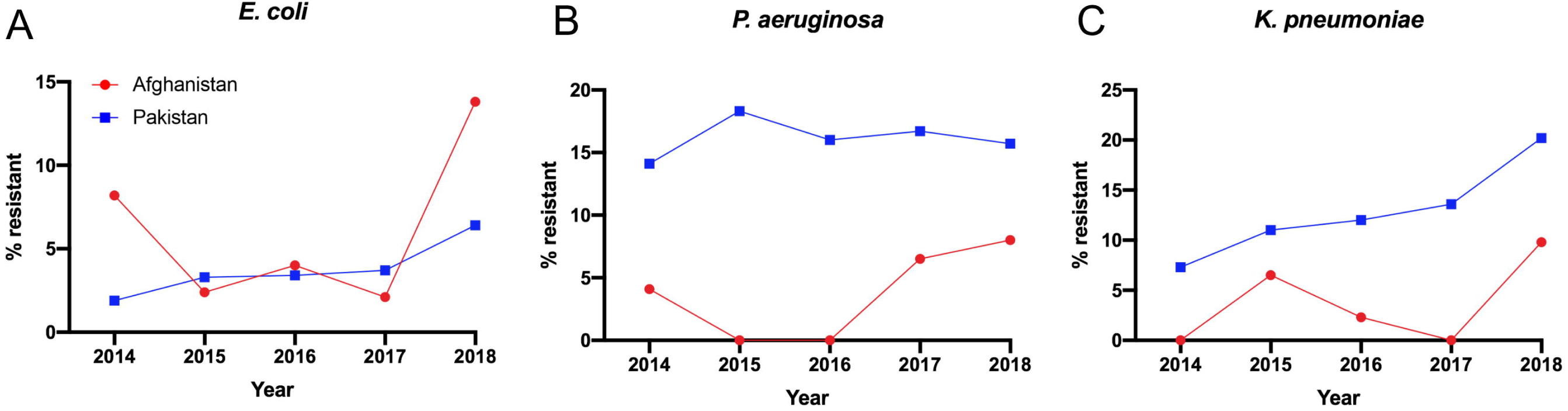
Trends in imipenem resistance in Adult patients, 2014-2018. Trends in resistance to imipenem for (A) *E. coli*, (B) *P. aeruginosa* and (C) *K. pneumoniae* isolates from adult populations with nationality from Pakistan or Afghanistan taken from SKCM&RC, from 2014-2018

**Figure 4.**
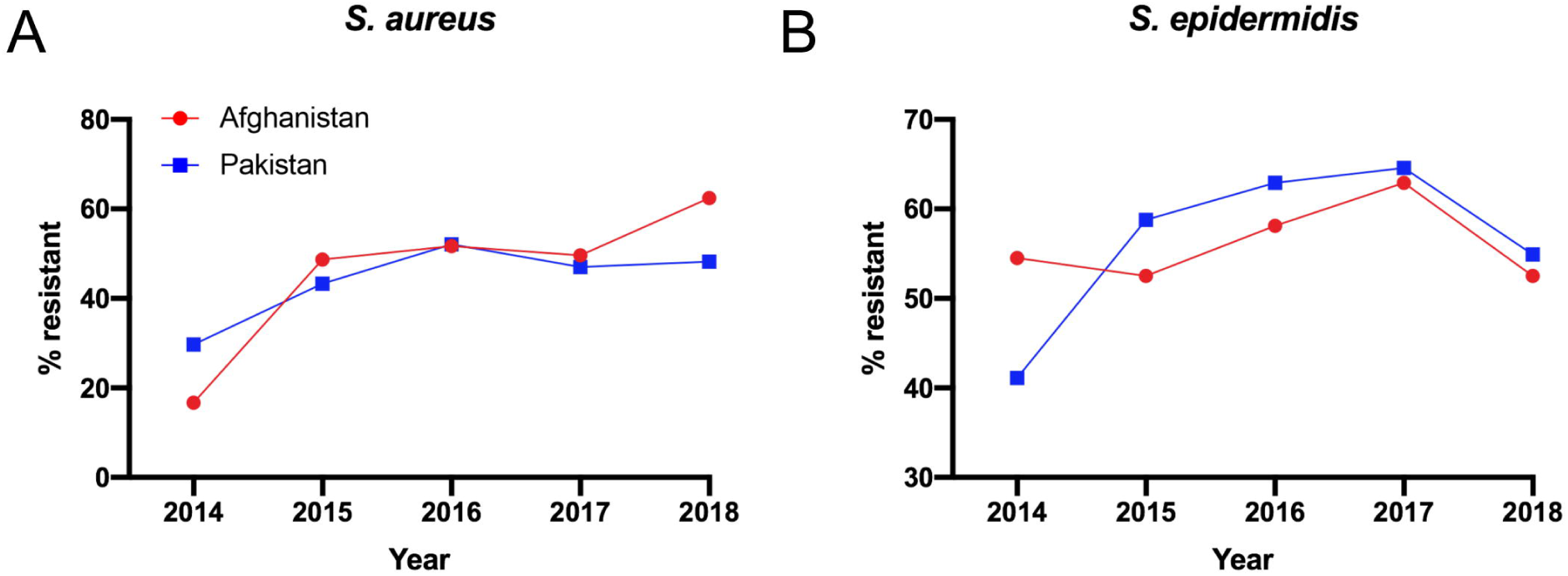
Trends in oxacillin resistance in Adult patients, 2014-2018. Trends in ciprofloxacin resistance for (A) *S. aureus*, and (C) *S. epidermidis* isolates from adult populations with nationality from Pakistan or Afghanistan taken from SKCM&RC, from 2014-2018

Another important last-line antibiotic is colistin. Colistin resistance for *E. coli, S. aureus, P. aeruginosa* and *K. pneumoniae* was detected at <1% for adults and children in Pakistan with the exception of *K. pneumoniae* in 2018. In this year colistin resistant *K. pneumoniae* for adults rose to 5.9% (n=222) and 2.2% for children (n=45) (Table S1-S4). It is important to track these types of changes to see if this is an emerging phenomenon. Colistin resistance was only detected in *P. aeruginosa* isolates at 1.4% (n=74) in 2016 in the adult population from Afghanistan.

### 3.5 Comparison to WHO GLASS reporting

Data available from the WHO GLASS for 2017 as reported by the Center for Disease Dynamics & Policy ResistanceMap[11] was compared to resistance rates reported here[9]. It should be noted that both Pakistan and Afghanistan are enrolled in GLASS. There is currently no reported AMR data for Afghanistan and data from 6 surveillance sites (4 hospitals and 2 outpatient facilities) from the 2018 call for data for Pakistan. For *S. aureus*, oxacillin for both populations levels were slightly below the confidence interval from GLASS for 2017 (63% (56-70%)), Table 4. For *E. coli* for both populations, ciprofloxacin resistance was ∼15% higher outside the confidence interval (56-62%) from GLASS. Carbapenem resistance was lower by ∼5% lower than the confidence interval reported (8-12%) (Table 5). Ampicillin and ceftriaxone were also just below confidence intervals reported (Table 5). For *K. pneumoniae*, ciprofloxacin resistance for the Pakistan population fell within GLASS values while that of the Afghanistan population was lower. For both carbapenems and third generation cephalosporins, % resistance for both populations were lower than reported by GLASS (Table 5). Our sample size, especially for the Pakistani population, is between 5.5 to 13-fold greater than the number of isolates that make up the resistance rates in GLASS.

**Table 5.** Comparison of resistance rates to GLASS averages for 2017.

|  | Antibiotic |  | Pakistan<br>(GLASS) |  | Antibiotic | Pakistan<br>(SKMCH&RC) |  | Afghanistan<br>(SKMCH&RC) |  |
| --- | --- | --- | --- | --- | --- | --- | --- | --- | --- |
|  |  |  | % resistant<br>(average<br>(95% CI)) | n<sup>1</sup> |  | % resistant | n | % resistant | n |
| *S. aureus* | Oxacillin | 2017 | 63 (56-70) | 193 | Oxacillin | 47 | 2390 | 49.6 | 115 |
| *E. coli* | Aminopenicillins | 2017 | 97 (95-98) | 427 | Ampicillin | 91.4 | 5566 | 93.8 | 96 |
|  | Fluoroquinolones | 2017 | 59(56-62) | 866 | Ciprofloxacin | 79.5 | 5578 | 77.6 | 98 |
|  | Carbapenems | 2017 | 10(8-12) | 814 | Imipenem | 3.7 | 5528 | 2.1 | 97 |
|  | Third Generation<br>Cephalosporins | 2017 | 86(83-89) | 458 | Ceftriaxone | 79.5 | 5290 | 86.7 | 98 |
| *K. pneumoniae* | Fluoroquinolones | 2017 | 58(51-65) | 189 | Ciprofloxacin | 56.2 | 1280 | 24.1 | 29 |
|  | Carbapenems | 2017 | 43(36-50) | 181 | Imipenem | 13.6 | 1275 | 0 | 29 |
|  | Third Generation<br>Cephalosporins | 2017 | 84(79-88) | 227 | Ceftriaxone | 65.3 | 1290 | 48.3 | 29 |

<sup>1</sup>The n beside each antibiotic represents the number of isolates that were tested for that antibiotic and which the % resistant was calculated from.

## 4. Discussion

Here, we present a situational report on antimicrobial resistance from a hospital and diagnostic center in Lahore, Pakistan. We find that the resistance rates largely are within or close to the range of previously reported values with a few notable differences (Table 5). However, national reporting for all antibiotics and bacteria are still sparse and increased reporting would provide a more comprehensive comparison. Notably, there were no striking difference in AMR rates for the Afghan population compared to the Pakistani population. As discussed early, a recent systematic review on AMR among migrant groups in Europe found AMR was higher in refugees and asylum seekers than other migrant groups, but did not find evidence of high rates of transmission of AMR from migrant to host populations [6]. Differences have been shown in the type of AMR between migrant and host populations. For example a study comparing German nationals to refugees shows differences in microbiome and prevalence of resistance genes [12]. A study of AMR in *Vibrio cholera* stains from Afghan patients in Iran showed differences in resistance patterns. In Afghan patients resistance to erythromycin, sulfamethoxazole trimethoprim and ampicillin was prevalent. In Iranian patients, resistance to tetracycline and nalidixic acid was enriched, however the sample size for this study were low[13]. We do not have genetic information to determine the underlying mechanisms of resistance.

Overall, this report agrees with previously reported levels of antibiotic resistance in Pakistan and does not demonstrate any strong differences among the AMR burden between national populations from one hospital and research facility in Lahore, Pakistan that provides diagnostic testing for locations around the county. This report, though, represents a specific clinical situation and more standardized reporting is needed to generate more robust country-wide estimates. Retrospectively, we observe the emergence of ceftriaxone resistant *S. typhi* (Table 3), however real-time analysis is critical for preventing outbreaks as they first emerge. National averages and baselines are important for hospitals to measure incidence of resistance and note any unusual changes or up or downward trends.

## Data Availability

Data is available upon request

## Ethical approval

Boston University Institutional Review Board oversight was not required as research was deemed not Human Subject Research. Exempt status for study was granted from the SKMCH&RC Institutional Review Board.

## References

[1] World Bank. Pakistan n.d. https://data.worldbank.org/country/pakistan (accessed October 31, 2019).

[2] Ministry of National Health Services Regulations & Coordination Government of Pakistan. National AMR Action Plan for Pakistan 2017:1–64.

[3] Saleem Z, Hassali MA, Hashmi FK. Pakistan’s national action plan for antimicrobial resistance: translating ideas into reality. Lancet Infect Dis 2018;18:1066–7. https://doi.org/10.1016/S1473-3099(18)30516-4.

[4] Global Antibiotic Resistance Partnership. Situational Analysis Report on Antimicrobial Resistance in Pakistan. 2018. https://doi.org/10.1017/CBO9781107415324.004.

[5] United Nations High Commissioner for Refugees. Pakistan n.d. https://www.unhcr.org/en-us/pakistan.html (accessed October 31, 2019).

[6] Nellums LB, Thompson H, Holmes A, Castro-Sánchez E, Otter JA, Norredam M, et al. Antimicrobial resistance among migrants in Europe: a systematic review and meta-analysis. Lancet Infect Dis 2018;18:796–811. https://doi.org/10.1016/S1473-3099(18)30219-6.

[7] Rice LB. Federal funding for the study of antimicrobial resistance in nosocomial pathogens: no ESKAPE. J Infect Dis 2008;197:1079–81. https://doi.org/10.1086/533452.

[8] Hasan B, Perveen K, Olsen B, Zahra R. Emergence of carbapenem-resistant Acinetobacter baumannii in hospitals in Pakistan. J Med Microbiol 2013;63:50–5. https://doi.org/10.1099/jmm.0.063925-0.

[9] World Health Organization. Global Antimicrobial Resistance Surveillance System (GLASS) Report. Geneva, World Health Organization. 2017. https://doi.org/ISBN978-92-4-151344-9.

[10] Klemm EJ, Shakoor S, Page AJ, Qamar FN, Judge K, Saeed DK, et al. Emergence of an extensively drug-resistant Salmonella enterica serovar typhi clone harboring a promiscuous plasmid encoding resistance to fluoroquinolones and third-generation cephalosporins. MBio 2018;9:1–10. https://doi.org/10.1128/mBio.00105-18.

[11] The Center for Disease Dynamics Economics & Policy. ResistanceMap:Pakistan 2018. https://resistancemap.cddep.org/CountryPage.php?countryId=85&country=Pakistan+ (accessed October 31, 2019).

[12] Häsler R, Kautz C, Rehman A, Podschun R, Gassling V, Brzoska P, et al. The antibiotic resistome and microbiota landscape of refugees from Syria, Iraq and Afghanistan in Germany. Microbiome 2018;6:1–11. https://doi.org/10.1186/s40168-018-0414-7.

[13] Tabatabaei SM, Salimi Khorashad A. Antimicrobial Resistance Patterns of Vibrio cholera Strains Isolated From Afghan and Iranian Patients in Iran. Int J Infect 2015;2:5–10. https://doi.org/10.17795/iji-22822.

